# The role of receptor binding and immunity in SARS-CoV-2 fitness landscape: a modeling study

**DOI:** 10.1101/2024.10.24.24316028

**Authors:** Zhaojun Ding, Hsiang-Yu Yuan

**Affiliations:** Department of Biomedical Sciences, College of Biomedicine, City University of Hong Kong, Hong Kong; Centre for Applied One Health Research and Policy Advice, Jockey Club College of Veterinary Medicine and Life Sciences, City University of Hong Kong, Hong Kong

**Keywords:** ACE2 binding score, Immune escape, Deep mutational scanning data, Rugged fitness landscape, SARS-CoV-2 evolution trajectory

## Abstract

Despite extensive research on SARS-CoV-2 ACE2 binding and transmissibility, their relationship concerning varying immunity remains unclear. SARS-CoV-2 RBD sequences from Italy in GISAID, combined with multiple deep mutational scanning data, were used to calculate ACE2 binding score and immune escape during the pandemic. We developed a COVID-19 transmission model that decomposed the effective reproduction number into three time-varying components: viral infectiousness (representing fitness), host susceptibility, and contact rate. After model fitting, a rugged fitness landscape, spanned by ACE2 binding score and virus-perceived effective immunity (adjusted for viral immune escape), was observed with peaks corresponding to individual VOCs (alpha, delta, and omicron (BA.1* and BA.2*)). Increasing effective immunity with reduced ACE2 binding corresponded to decreasing virus fitness peaks from alpha to delta. Among omicron sub-lineages, which exhibited enhanced immune escape, BA.2* reached a fitness optimum while retaining slightly reduced ACE2 binding relative to BA.1.1. The findings help explain the evolution of SARS-CoV-2.

## INTRODUCTION

The COVID-19 pandemic was characterized by repeated infection waves, driven by SARS-CoV-2 variants with new and advantageous genomic mutations. These mutations enhanced fitness in variants of concern (VOC) via a complex interplay between viral intrinsic traits and population immunity, shaped by vaccinations and prior infections ^1,2^. Therefore, understanding the SARS-CoV-2 fitness landscape can provide insights into evolutionary virus dynamics, knowing whether the virus undergoes gradual or punctuated evolution, which is important for predicting and preparing for future outbreaks.

The SARS-CoV-2 fitness is associated with virus’s ability to bind to ACE2 receptors and evade immune defenses^1^. Upon entering the human body, the virus uses its spike protein to bind to the ACE2 receptor on the surface of host cells. This interaction occurs primarily through the receptor-binding domain (RBD). Simultaneously, the host immune system produces neutralizing antibodies, which prevent the virus from binding to ACE2. However, SARS-CoV-2 can evade immune recognition by mutating amino acid residues within the RBD, a phenomenon known as immune escape ^1,3^. However, how ACE2 binding and immune escape interact to influence virus fitness under varying immunity levels among infected hosts remains unclear.

The accumulated mutations in the RBD of SARS-CoV-2 spike protein can confer selective advantages toward a higher fitness, resulting in antigenic drift ^2^. For example, the N501Y mutation in the alpha (B.1.1.7) sub-lineage may have altered spike protein binding affinity to ACE2, resulting in higher transmission rates ^4,5^. This mutation appeared in the omicron sub-lineages. Such evolutionary changes help sustain the virus’s presence in the population, facilitating COVID-19’s transition from a pandemic to an endemic state characterized by periodic surges.

The fitness landscape has been developed to represent the relationships between genetic variations and their corresponding fitness values, providing insights into the potential adaptive trajectories of populations ^2,6^. A rugged landscape occurs due to interactions between mutations (epistasis) ^6,7^. Increasing immunity can also exert high selection pressure on viruses, contributing to antigenic drift ^2,8^. When a virus evolves through fitness valleys toward a new peak, it generates boom-and-bust patterns, likely resulting in repeated outbreaks ^9^. However, when SARS-CoV-2 evolutionary and transmission dynamics are intertwined, reconstructing the fitness landscape becomes challenging. For example, not only virus traits, but also host immunity (e.g. vaccination) and host behaviors (e.g. social distancing measures and public awareness) can influence transmissibility (or effective reproduction number) ^10^. Hence, quantifying viral fitness and finding its relationship with viral traits presents a challenge.

In this study, we aimed to assess the relationship between viral fitness and traits derived from deep mutational scanning (DMS) during the pandemic. Sequence and epidemiological data from Italy, a country with high viral incidence rates and high vaccine coverage, were used. To quantify and thereby separate intervention effects (i.e., vaccinations and non-pharmaceutical interventions (NPIs)) on virus fitness, we incorporated mobility and vaccination data in an infectious disease transmission model to calibrate fitness estimates. From simulated results, we constructed a fitness landscape after mapping the virus’s evolutionary trajectory traits (i.e., ACE2 additive sum of binding score and immune escape) over different periods across different immunity levels. This study was previously made available as a preprint version ^11^.

## METHOD DETAILS

### Sequence data

Italian SARS-CoV-2 spike protein sequence data (31^st^ January 2020–17^th^ October 2022) were downloaded from the GISAID database ^12^. To ensure data quality control, two exclusion criteria were applied: (1) sequences with > 5% ambiguous amino acids ^13^ and (2) sequences < 1,263 bp or > 1,273 bp were filtered out. Ultimately, 112,011 sequences were included for virus ACE2 receptor binding and immune escape measurements. The proportion of beta and gamma sequences were very low, only 2.07% and 0.12%, respectively (see Supplementary Fig. S1). Therefore, only alpha, delta, and omicron variants were reported as the main VOCs in subsequent analyses.

### Measuring virus ACE2 additive sum of binding score and immune escape

DMS data were used to calculate virus ACE2 additive sum of binding score (i.e., the sum of individual mutational effects) by mapping binding affinities between ACE2 and SARS-CoV-2 RBD amino acid mutation sites ^14^. To reduce the confounding effects of epistasis in cell binding, we employed multiple DMS datasets generated from five variants as backgrounds, namely Wuhan-Hu-1, Alpha, Delta, BA.1, and BA.2 ^15,16^. For each viral sequence, we selected the DMS dataset whose reference background most closely matched the sequence’s ancestral lineage and used it to compute the ACE2 additive binding score (e.g., wild-type → Wuhan-Hu-1; Alpha and its sub-lineages → Alpha; Delta and its sub-lineages → Delta; BA.1 and its sub-lineages → BA.1; BA.2 and its sub-lineages → BA.2).

Escape fraction data were used to determine viral immune escape by mapping reduced binding levels between the virus and antibodies, using datasets integrated from 33 monoclonal antibodies and polyclonal sera, as available from the SARS-CoV-2 RBD antibody escape maps (https://jbloomlab.github.io/SARS2_RBD_Ab_escape_maps/) ^17^. First, we used the Clustal Omega tool to align all sequences ^18^. Next, to calculate virus ACE2 additive sum of binding score for each sequence, binding changes across individual amino acid mutations of RBD region (residues from N331 to T531) were summed (see Supplementary Fig. S8) ^19^. For immune escape, we calculated the mean reduction in antibody binding for each amino acid mutation site across 33 monoclonal antibodies. Immune escape across individual amino acid mutations was summed, like the calculation of ACE2 binding. Finally, we computed daily average values of the additive ACE2 binding score and the additive immune escape score among all daily samples. Further details are shown in Supplementary Methods 2.1.

### Mobility data and calculating the mobility index

The mobility index, which indicated the degree of population contact, was estimated from Google Mobility and Apple Mobility data ^20,21^. Specifically, three Apple mobility data subtypes (driving, walking, and transit movement) and four Google subtypes (grocery and pharmacy stores, transit, retail and recreation, and workplaces) were used to calculate mobility indices based on previous research ^22^.

### Vaccination data

COVID-19 vaccine administration data provided by the Italian Ministry of Health were used to summarize daily vaccination numbers for first, second, and third doses ^23^. During the study period (i.e., model fitting period, 31^st^ January 2020–5^th^ March 2022), 134,611,131 vaccine doses were recorded and sourced from five major vaccine manufacturers: Janssen, Moderna, Novavax, Pfizer, and AstraZeneca (see Supplementary Table S1). Messenger RNA (mRNA) vaccines (i.e., Pfizer and Moderna) and viral vector vaccines (i.e., Janssen and AstraZeneca) accounted for almost 99% of the Italian vaccination campaign (see Supplementary Fig. S3). Consequently, we only considered mRNA (Pfizer and Moderna) and viral vector vaccines (Janssen and AstraZeneca).

### Calculating virus fitness

A modeling approach was adopted to quantify viral infectiousness, which we refer to as viral fitness throughout this study. We extended a stochastic Susceptible - Exposed - Infectious - Recovered (SEIR) model, coupled with vaccination, mobility, and immune escape data, to reproduce COVID-19 transmission dynamics and estimate virus fitness across different VOC periods (see Fig. 1A, B). The model was used to estimate the part of the transmission rate that were determined by viral ACE2 receptor binding only over time (denoted as *β_v_(t)*) under different immunity levels, representing the change of infectiousness. The relative virus fitness was defined as *β_v_(t)/β_0_*, where *β*_0_ is the average value of *β_v_(t)* during the first month of the pandemic, representing the wild-type.

**Fig. 1.**
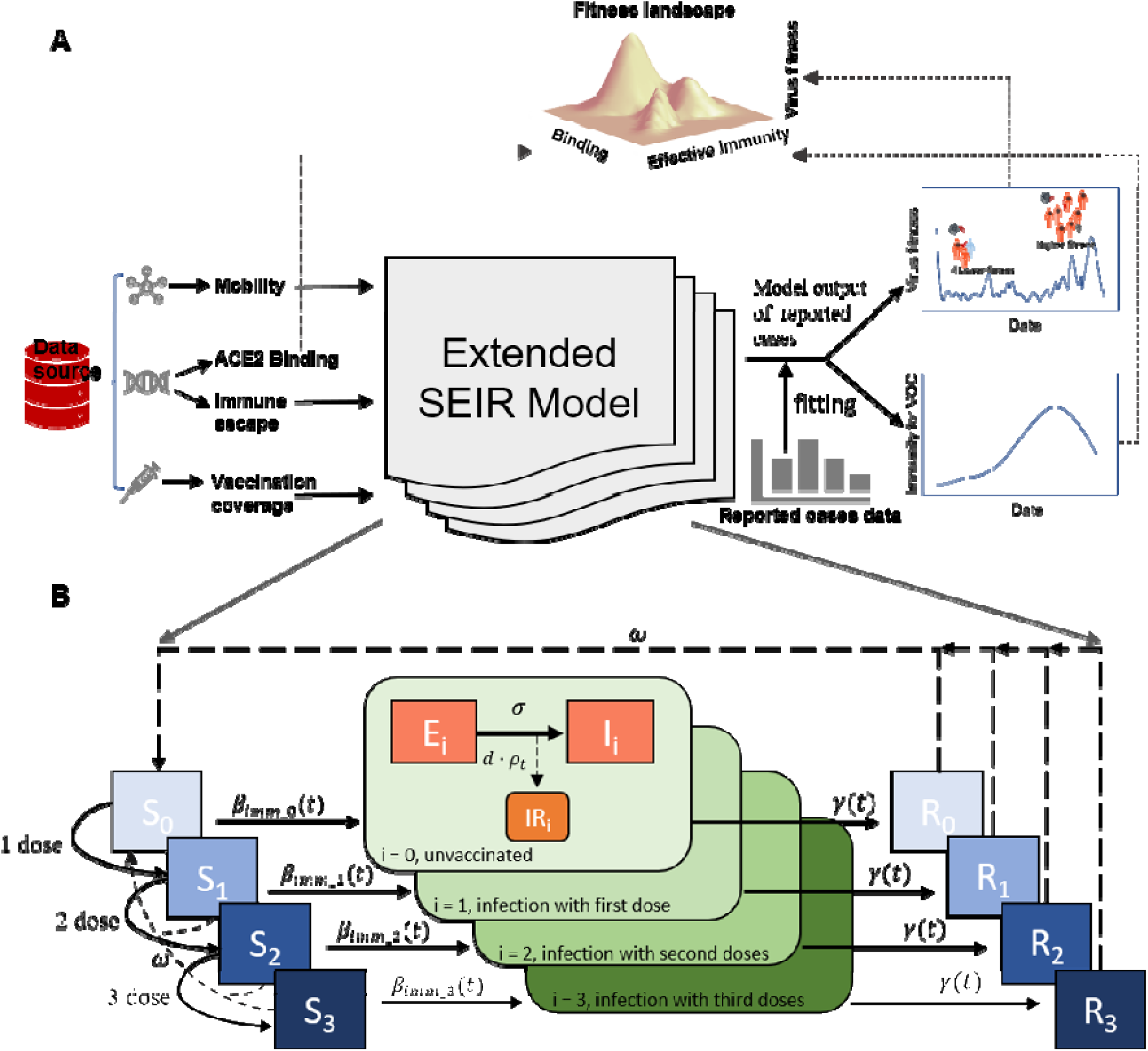
Study flow and model design. (A) Schematic showing the extended SEIR model integrating multiple data sources to estimate the SARS-CoV-2 fitness landscape. Data inputs include mobility, immune escape, and vaccination coverage data. These data feed into the extended SEIR model, which was calibrated using reported case data. The model outputs include virus fitness combined with ACE2 binding data and effective immunity, and show how these factors shape virus evolution patterns. (B) Detailed structure of the SEIR compartment model. The model partitions the population based on vaccination status and infection stage, highlighting transitions between susceptible (S), exposed (E), infectious (I), reported infectious cases (IR) and recovered (R) states, with an emphasis on the impact of vaccination doses on these dynamics. S_i_, E_i_, I_i_, IR_i_ and R_i_ subscripts denote compartments with different vaccine doses.

The effective reproduction number (R_e_) was represented as two components (see Fig. 2D): an intrinsic factor (infectiousness), and extrinsic factor (susceptibility and contact rate), as below.

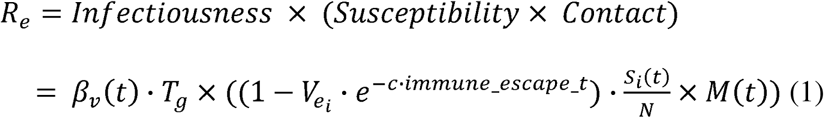

**Fig. 2.**
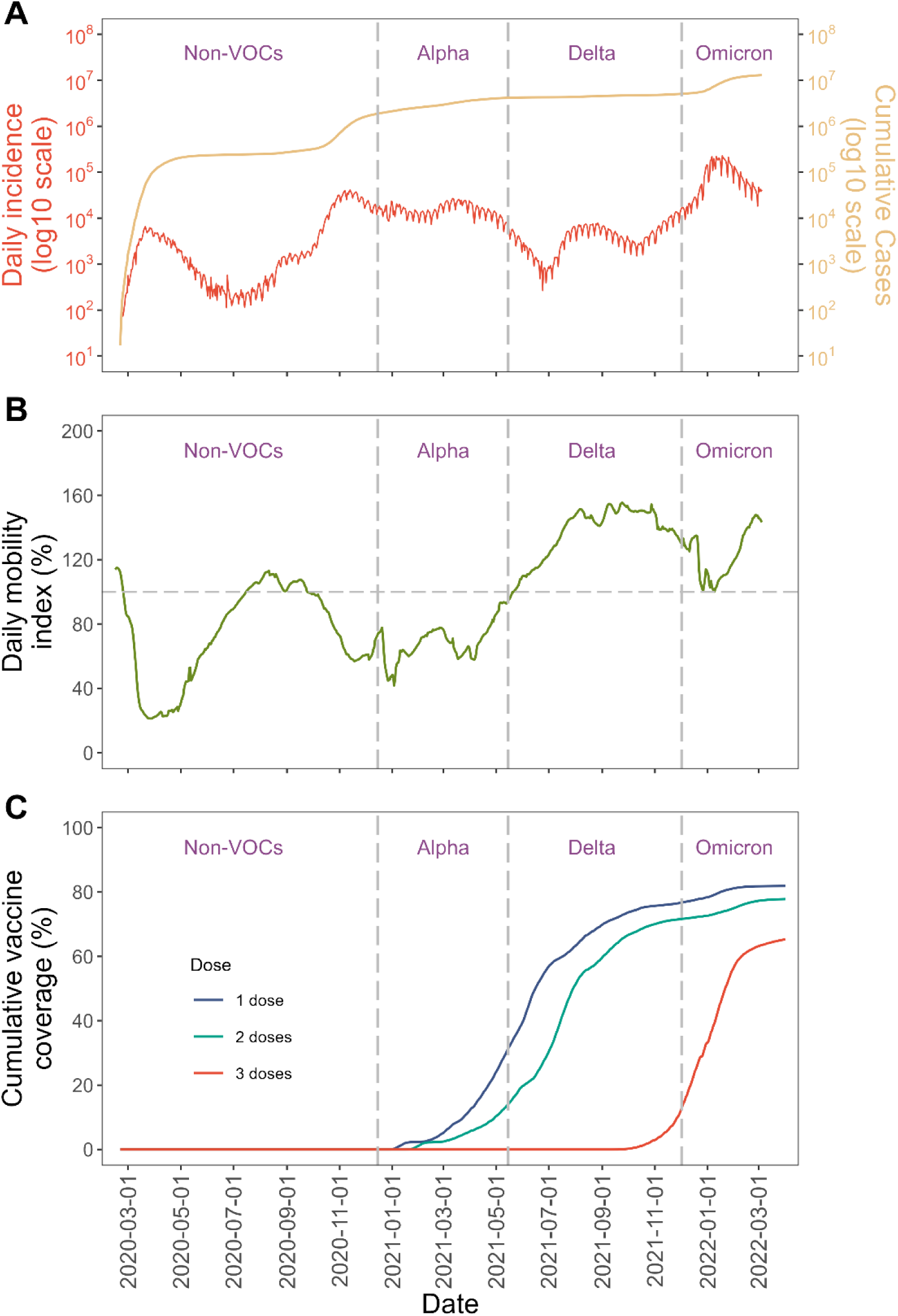
Factors influencing daily COVID-19 incidences and virus fitness. (A) Daily COVID-19 incidences in Italy. New daily COVID-19 cases in Italy are measured in log10 scales. VOC emergence is highlighted by vertical dashed lines, labeled as non-VOC, alpha, delta, and omicron strains. (B) Vaccination coverage in Italy from the first to the third vaccine doses (C) Mobility index in Italy. Population movement levels relative to pre-pandemic levels (horizontal dashed line) are shown.

where T_g_ is the generation time, which is 4 days^22–26^; *V_ei_* (t) refers to vaccine efficacy from different doses (*i*) against the wild-type; *V_ei_* ·e^-c·immune_escape(t)^ represents protection against circulating strains, accounting for their immune escape. Immune_escape_t is the daily average immune escape of viruses; c is the scaling factor for daily immune escape; 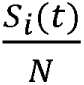 represents the fraction of individuals at each susceptibility level (i.e., S_0_, S_1_, S_2_ or S_3_), based on vaccine dose *i* received; M(t) is the daily mobility index, representing the effect of the change in contact rate on reducing overall fitness, 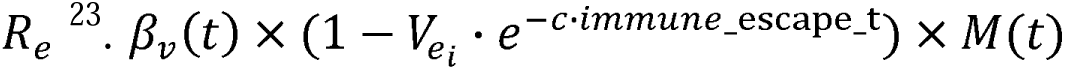 represents the transmission rate among susceptible groups with different vaccine efficacy (i.e., β_imm_i_(t) in Fig. 1B). The effective reproduction number R_e_ was obtained after fitting the transmission model (Fig. 1B) to the daily reported cases, when daily vaccine doses were considered.

Exposed (E_i_) individuals who came into contact with infected individuals will become infectious 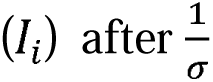 days. As not all cases were reported correctly and immediately, reported infectious cases (IR) were considered based on the detection rate (d) and the confirmation delay (p_t_) in the transition between E_i_ and I_i_, which was used to fit the daily reported cases. Individuals with an infectious status would subsequently enter a removed status (i.e., R_0_, R_1_, R_2_ or R_3_) due to self-isolation or recovery after 1/γ(t) days. Considering waning immunity among these individuals, we assumed that recovered and vaccinated individuals lose their immunity on average after ω days (ω = log(0.85)/182.5, corresponding to exponential waning with a 15% loss in protection after 6 months), which is the same to the studies by Barnard et al. and Lau et al.’s settings ^27,28^. In addition, we also used a fast vaccine waning rate (ω = log(0.5)/90, 50% loss of protection after 3 months) and moderate waning rate (ω = log(0.5)/182.5, 50% loss of protection after 6 months) as the sensitivity analysis to check the stability of fitness patterns under different waning scenarios.

### Population immunity and effective immunity

Population immunity (P_imm_total_) refers to the average level of immunity to a specific infectious agent (e.g., Wuhan-Hu-1) within a population, achieved through both natural infection and vaccination. It is commonly expressed as the proportion of individuals in the population who are immune, either through previous infection or through vaccination, but partial protection and waning immunity should be considered ^29,30^. These factors were incorporated into our analysis. We further calculated the virus-perceived immunity, termed effective immunity (P_imm_eff_) as P_imm_eff_ = P_imm_total_ (t) · e^-c·immune_escape_t^, which accounts for immune escape and represents protection against currently circulating strains. More details about the parameters set and population immunity calculations can be found in the Supplementary Methods 2.2.

### Model fitting for reproduce of infection dynamics and fitness landscape constructed

The number of individuals in compartment IR (i.e., reported infectious cases), as the modeling output, was used to fit the daily numbers of reported cases. The Particle Markov chain Monte Carlo method ^31,32^, which combines particle filtering and Markov chain Monte Carlo approaches, was used to calibrate model output using daily reported cases and then estimate model parameters, with a total of 100,000 iterations. All model parameter estimations were obtained using the ‘mcstate’ package in R ^31^. A fitness landscape was fitted with virus fitness and assumed a multivariate normal distribution, which helped visualize SARS-CoV-2 adaptive evolutionary trajectories. More technical details about fitness landscape reconstruction can be found in the Supplementary Methods 2.2.4.

## RESULTS

### The relationship between daily mobility, vaccine coverage, and daily incidence

A large variety in daily incidence and mobility was observed (Fig. 2A, C and Supplementary Fig. S2). Despite ongoing vaccination efforts (Fig. 2B), viral outbreaks reoccurred and cumulative cases increased rapidly during the omicron period (Fig. 2A). The effective reproduction number was likely influenced by both virus infectiousness and susceptibility. In our model, we considered susceptibility (determined by population immunity and immune escape) and contact rate (determined by mobility) as extrinsic factors while virus infectiousness as an intrinsic factor (i.e., viral fitness; see Methods for the definition).

### Temporal dynamics of virus ACE2 additive sum of binding score and immune escape

We observed distinct patterns between the two viral traits across main VOCs, including alpha, delta, and omicron (Fig. 3A). ACE2 additive binding score decreased from a high level in alpha (B.1.1.7) to delta (B.1.617.2), with early omicron lineages (BA.1*) showing binding scores comparable to the wild-type (Fig. 3B). Binding scores then slightly fluctuated among omicron sub-lineages, with BA.1.1 showing an increase relative to BA.1, and BA.1# showing a decrease.

**Fig. 3.**
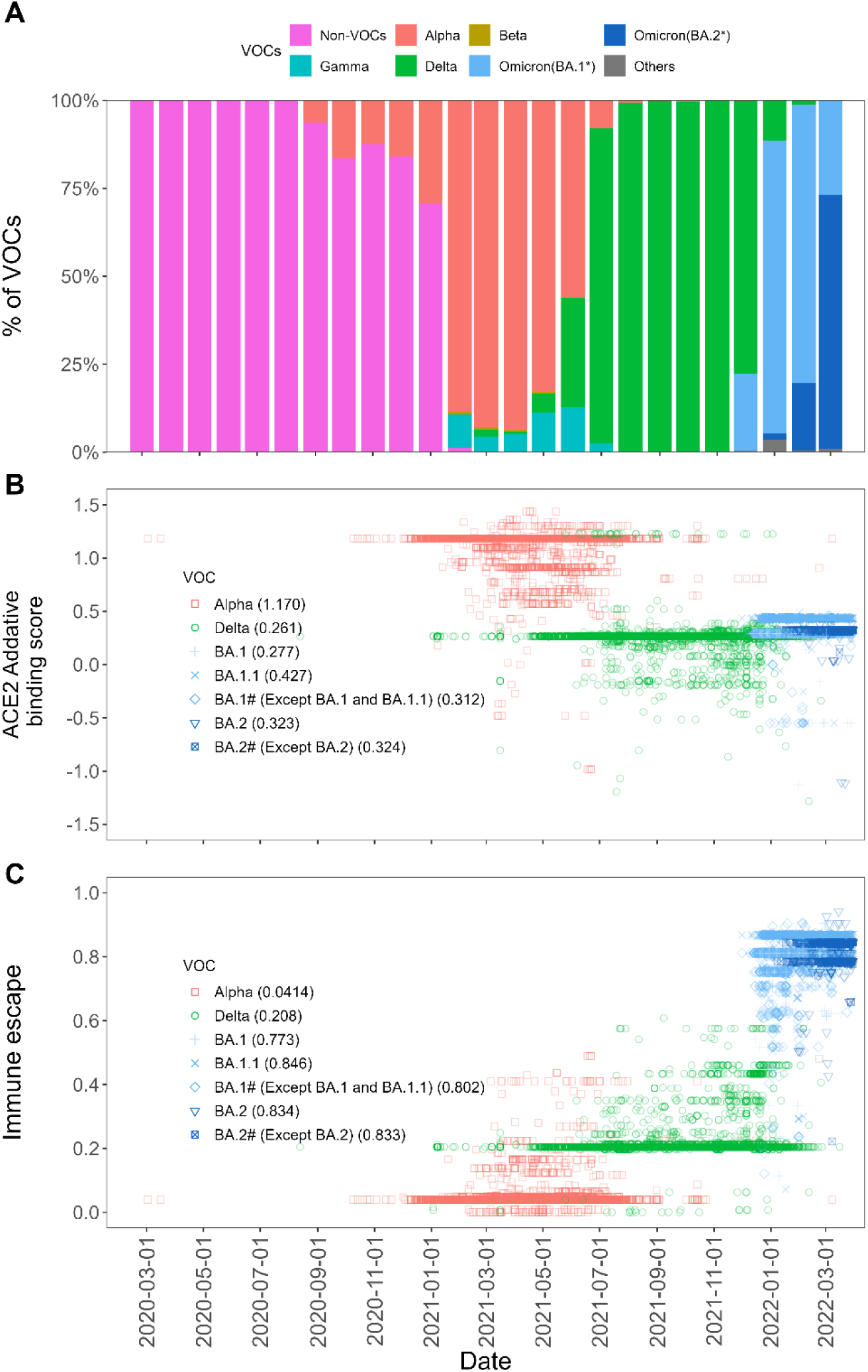
Comprehensive analysis of SARS-CoV-2 VOC with ACE2 binding affinity and immune escape. (A) Variant prevalence over time. The bar chart shows the prevalence of different SARS-CoV-2 VOC (alpha, beta, delta, gamma, and omicron strains) from early 2020 through early 2022. (B) Additive binding scores of viral lineages under different VOC backgrounds. The value in parentheses after each VOC denotes the mean ACE2 binding scores. (C) The immune escape scores over time. Scatter plot showing ACE2 binding and alpha, delta, and omicron variant immune escape across the timeline. BA.1* and BA.2* including BA.1, BA.2 and their sub-lineages. BA.1# means BA.1* without BA.1 and BA.1.1. BA.2# means BA.2* without BA.2.

The emerging BA.2* lineages subsequently showed ACE2 binding scores between BA.1 and BA.1.1. On the other hand, immune escape showed a gradual rise from alpha to delta, followed by a sharp rise with the emergence of omicron, whose sub-lineages exhibited similar levels. (Fig. 3C).

### SARS-CoV-2 transmission dynamics

To understand how ACE2 receptor binding might influence virus fitness, we incorporated vaccine coverage, mobility and virus immune escape data into the model to calibrate model fitting for daily reported cases (Fig. 1B). Our model was able to reproduce the observed transmission dynamics, including Non-VOC and VOC-associated epidemic waves (Fig. 4A). The relative virus fitness was increasing form late delta to omicron periods (Fig. 4B). Both vaccine-induced immunity (Fig. 4C, green line) and total natural infection combined with vaccine-induced immunity (Fig. 4C, red line)) increased due to the initiation of the vaccination campaign (beginning in the late December 2020) in Italy around the same time ^33^. A similar trend was observed for effective immunity (i.e., immunity which considering the waning immunity and immune escape of circulating VOCs) against VOCs during alpha and delta periods, but a significant decrease was recorded in the omicron period (Fig. 4D). This decline reflects the strong immune escape capabilities of omicron, despite the ongoing vaccination campaign.

**Fig. 4.**
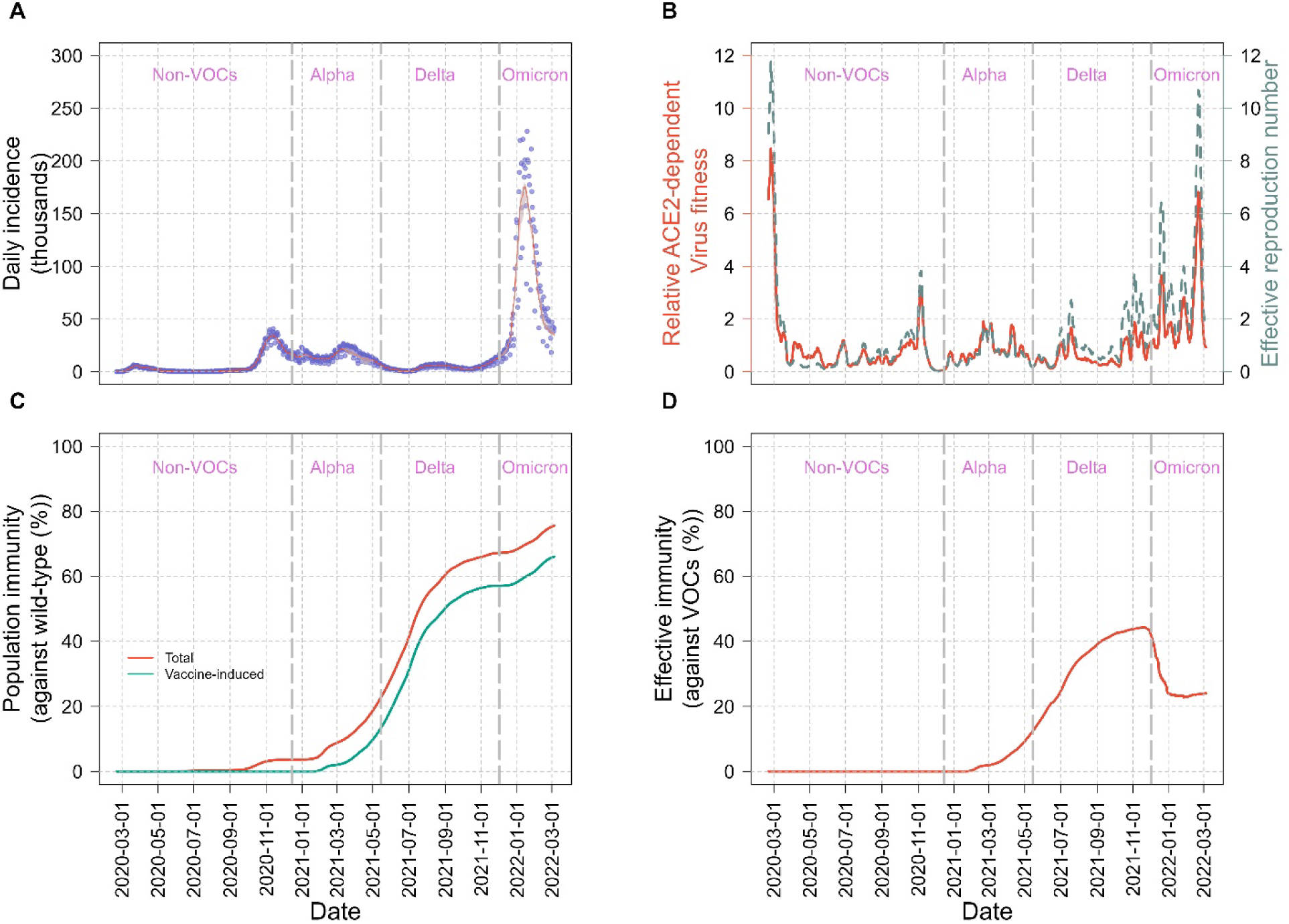
Epidemic indicator and immunological response trajectories during SARS-CoV-2 variant transition in Italy. (A) Model fitting for daily confirmed cases. The red line represents the estimated daily incidences. (B) Virus fitness and effective reproduction number. The red line shows estimated virus fitness over time, which is relative to the wild-type virus. The dashed grey line shows effective reproduction number. (C) Population immunity against wild-type. The green curve represents the average proportion of the population protected by vaccine-induced immunity, while the red curve while the red curve reflects total immunity, including vaccine and infection-acquired immunity. (D) The curve represents the immune protection in the total population against circulating VOCs (referred to as effective immunity), considering virus immune escape properties.

### SARS-CoV-2’s fitness landscape

The phenotype-fitness landscape spanned by the additive sum of ACE2 binding score and host’ effective immunity showed rugged patterns with four major peaks, corresponding to B.1.1.7 (alpha), B.1.617.2 (delta), BA.1* (first omicron peak) and BA.2* (second omicron peak); One minor peak, corresponding to the transition period (mixed with delta and original omicron (B.1.529) lineages) between delta and omicron, and four valleys between these peaks. A higher effective immunity was associated with a lower fitness peak from alpha to delta. During the omicron period, a reduction in effective immunity from BA.1* to BA.2* together with similar ACE2 binding scores, corresponded with increasing fitness peaks (Fig. 5A). Notably, BA.2* maintained high immune escape scores comparable to BA.1.1, but showed slightly lower binding scores.

**Fig. 5.**
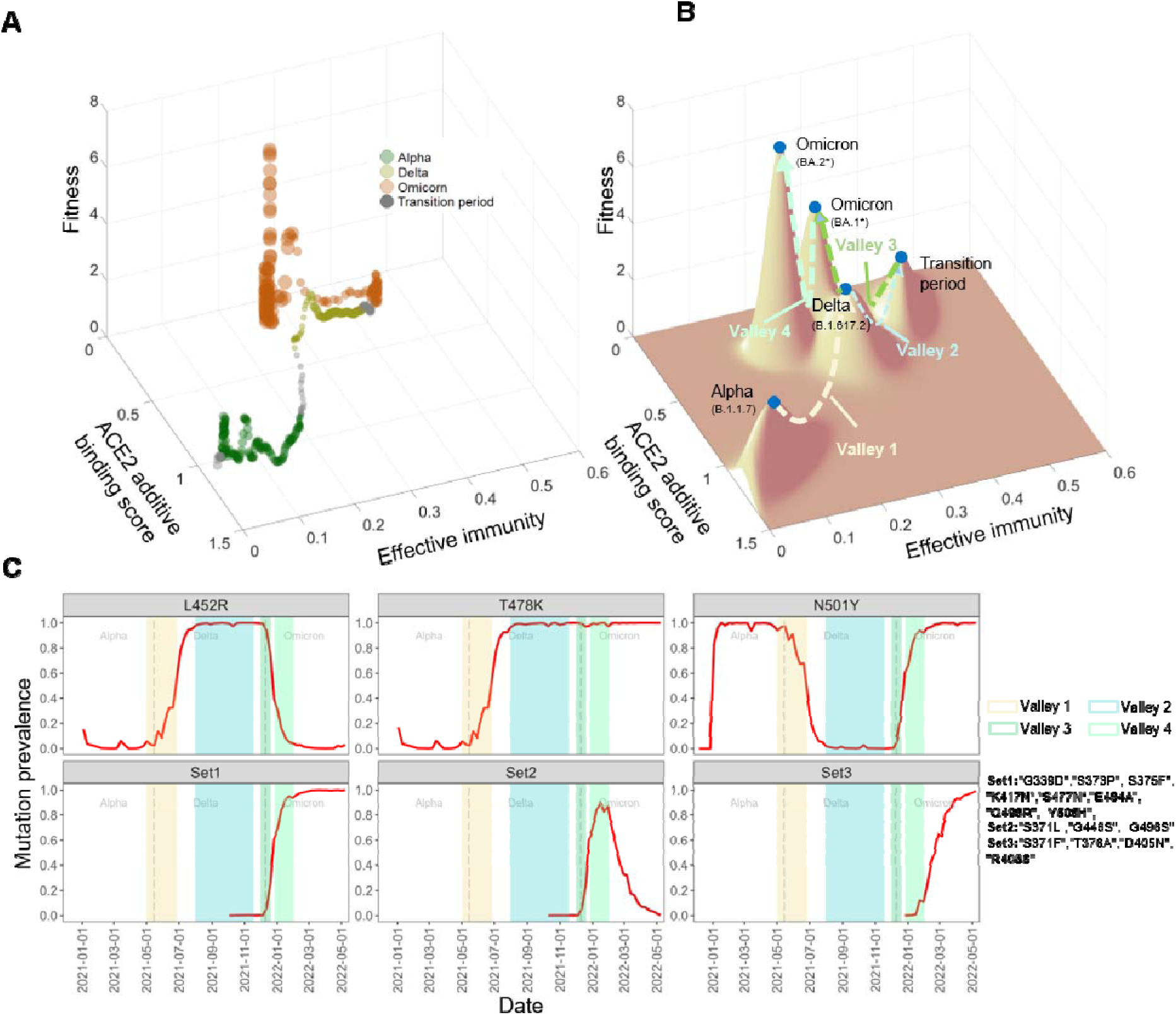
Interplay between ACE2 additive binding score, effective immunity, and virus’s fitness in SARS-CoV-2 evolution. (A) Observed relationship between ACE2 additive binding score, effective immunity and fitness. Gray dots represent variant fitness levels during a 2-week transition period following the initial outbreak of each VOC pandemic. (B) Fitness landscape of VOC. Surface topography shows the virus’s fitness landscape, which reconstructed based on multivariate normal distribution. Peaks denote maximum variant fitness values in predominant periods. (C) Mutation prevalence associated with VOC transitions. Shadowed regions corresponding to valleys in panel B are marked by different colors. Some cluster mutations with similar prevalence were termed “Set1”, “Set2” and “Set3”.

To evaluate robustness of the evolutionary pattern (i.e., Fig 5A), we varied vaccine waning from an optimistic baseline to moderate and fast scenarios (50% loss at 6 and 3 months, respectively, see supplementary Fig. S7). The shape and key features of the phenotype–fitness landscape was maintained, with no material shifts in variant ordering or trend direction. To help visualize the rugged patterns, a smoothed fitness landscape was constructed after fitting the fitness data (Fig. 5B; see Methods).

The sequential change in mutations, characterized by partial replacement, appeared through the fitness valleys toward the peaks were explored:

**Alpha -> delta:** During the transition between the fitness peaks at alpha and delta, the prevalence of N501Y (ACE2 binding changes (Wuhan-Hu-1 background, denoted as Δ_bind_): 0.96, immune escape changes (denoted as Δ_escape_): 0.01, see Supplementary Fig. S5), a mutation with high ACE2 binding, began to decline after passing through the first fitness valley (see Supplementary Fig. S5B and Fig. S6). Meanwhile, the prevalence of L452R (Δ_bind_: 0.16, Δ_escape_:0.18) and T478K (Δ_bind_: 0.04, Δescape:0.004) started to increase during this valley, gradually replacing N501Y as the dominant mutations. Some rare mutations, like E484K (Δ_bind_: 0.11, Δ_escape_:0.27), observed as fluctuation during the transition between the fitness peaks at alpha and delta. More details about the rare mutations across each VOC can be found in Supplementary Fig S5 and S6.

**Delta-> Omicron (BA.1*):** After the delta variant, the virus’s evolutionary pathway moved through two fitness valleys (i.e., Valley 2 and Valley 3) with a local peak (i.e., a minor peak during the transition period including a mixture of omicron and delta) and then reach to BA.1*, the next peak. Meanwhile, only N501Y and T478K was present in the BA.1* without L452R (Fig. 5C). Next, the Set 1 (G339D, S373P, S375F, K417N, S477N, E484A, Q498R and Y505H), Set 2 mutations (S371L, G446S and G496S) and few other mutations (N440K, Q493R and N501Y, see Supplementary Fig. S5) gradually increased as the BA.1 lineage predominated. During this period, effective immunity increased from around 0.2 to 0.4 and then returned to the original level, while binding values remained similar, corresponding to the suboptimal fitness.

**Omicron (BA.1*) -> Omicron (BA.2*):** The virus continued to evolve through Valley 4 towards the optimal fitness peak associated with BA.2*. During this transition, the Set 1 and other additional mutations (N440K, Q493R and N501Y) were retained while Set 2 gradually decreased as transient mutations. In the end, the Set 3 (S371F, T376A, D405N and R408S) gradually increased and became dominated. The mutational effects in Set 3 showed either unchanged or reduced binding scores using BA.1 as background (Supplementary Fig. S5). As the virus progressed toward BA.2 dominance, optimal fitness emerged, coinciding with reduced effective immunity and a moderate change in ACE2 binding (Fig. 5B).

## DISCUSSION

We introduced a mathematical framework to estimate virus fitness while reproducing the transmission dynamics of different SARS-CoV-2 variants in Italy. For the first time, the fitness landscape was constructed for the virus from alpha to omicron. This was achieved by incorporating both intrinsic viral factors (e.g., infectiousness) and extrinsic factors (e.g., host susceptibility and contact rates) into an SEIR-based model, which helps quantify virus fitness while removing the effects from extrinsic factors. The results provide valuable insights into the real-world evolutionary constraints of SARS-CoV-2. DMS data are used to provide a proxy of virus traits for sequences from GISAID. While ACE2 binding scores derived from DMS were used to characterize receptor affinity, additional binding results can be incorporated to further refine the fitness landscape.

Rugged fitness landscapes that were observed in our research (Fig. 5B and supplementary Fig. S7) suggest that certain factors may play important roles in the evolutionary pathway via fitness valleys, facilitating the transition to new fitness peaks. Epistasis can play a crucial role, as interactions between mutations are likely to lead to non-linear effects, forming rugged fitness landscapes, as we observed ^6,34,35^. Epistatic interactions remain uncertain as previous studies have reported conflicting findings on the trend of SARS-CoV-2 ACE2 receptor binding and the available experimental data are limited ^36–38^. Hence, we adopted multiple DMS datasets, each using a different variant or subvariant template. This potentially reduces the biases caused by epistatic effects more as the dissociation constant for each template sequence was directly measured rather than inferred by summing mutational effects from the wild-type ^15,16^.

Furthermore, the fitness valley maybe also related to deleterious effects among immune escape mutations ^39,40^. Those mutations occurred in the fitness valley may be detrimental to SARS-CoV-2’s evolution, requiring additional time to reach a fitness peak when compensatory mutations occur. In addition, recombination can be a potential driver of evolutionary change in SARS-CoV-2, especially when multiple variants co-circulate ^41^. This event can also result in viral fitness changes.

A possible compromise between receptor binding affinity and immune escape during RBD evolution of omicron sub-variants from BA.2 to BA.4/5 has been shown in a recent study ^42^. The authors found that a reduction of immune escape was compromised by a high cell binding. On the other hand, a strong immune escape mutation occurred with a reduced cell binding level. Early omicron evolution (from BA.1* to BA.2*) showed that once a suboptimal fitness peak is reached by strong immune escape, further adaptation occurred with fluctuating binding score. Furthermore, the constantly changing population immunity may play an important role in evolutionary shifts between different fitness peaks. In a previous study, the effects of immune escape mutations on viral fitness were compared between the presence and absence of antibodies, showing that these mutations can also affect replication ^43^. This highlights that fitness changes may also result from alterations in ACE2 binding. In our analysis, we considered immune escape to influence both host susceptibility and effective immunity. Consequently, changes in effective immunity modulated viral fitness through their interaction with ACE2 binding.

### Implications

Firstly, continuous surveillance efforts of SARS-CoV-2 is important. Even within the same VOC, such as omicron, virus still experienced rugged fitness landscape, leading to repeated epidemics of varying sizes. This is exemplified by the transition from omicron BA.1* to BA.2*, where the virus’s evolutionary path traversed a fitness valley before reaching a new peak. Knowing the change in traits will be important to understand why a new outbreak occur.

Secondly, forecasting when a large outbreak will occur is still very challenging. An optimal virus fitness was likely to be reached until a set of mutations having strong immune escape with required reduction in binding.

Moreover, the results indicate a complex relationship between virus fitness and host immunity, modulated by ACE2 binding. To better understand the complex interplay between multiple viral traits and fitness, more advanced artificial intelligence (AI) models, such as spiking neural networks (SNNs), may provide valuable insights. Such modeling approaches together with the phenotype-fitness landscape can help illuminate the dynamic evolutionary landscape of SARS-CoV-2, which in turn underscores the ongoing need for regular updates to vaccines to maintain their effectiveness against emerging variants.

### Study limitations

There are certain limitations in this study. Firstly, the study only focused on RBD region in the spike protein and we assumed the impact of each amino acid on viral traits is additive. Nonetheless, several experimental studies which consider the epistasis effect showed similar pattern in the order with our results of binding changes among each VOC ^37,44^. Secondly, the population mobility index was an NPI proxy as it assumed social contacts were proportional to population mobility, which not considered the awareness-driven behavior changes on the dynamic of mobility. There is a risk that changes in *β*_v_(t) may reflect changes in NPIs or awareness, which could cause the actual detection rate to vary over time and potentially influence the estimation of *β*_v_(t). Thirdly, our model categorized the population based on administered numbers of vaccine doses, without considering age and gender heterogeneity in the population. This simplification overlooked potential differences in vaccine responses and exposure risks across different demographic groups. Fourthly, we acknowledge that viral fitness may be influenced by within-host replication, survival, and other viral or host factors. While modeling simplifies the complexity, it allows us to understand the mechanism through quantifying and, therefore, separating individual effects. Fifthly, GISAID may contain some noisy data. We have done initial screening to remove data with apparent errors. Lastly, but critically, the seasonal or climatic factors were not considered. Future research should introduce the impact of these factors on virus fitness to provide a more comprehensive understanding of virus transmission dynamics.

## Conclusions

SARS-CoV-2 mutations along with immunity influences generated a rugged fitness landscape, which shaped viral evolutionary trajectories in Italy. Knowing the landscape can help provide a better understanding of the complex dynamics of viral evolution and adaptation, particularly in drug resistance and vaccine development. More importantly, host immune systems and viruses are constantly engaged in an arms race, with each adapting to changes in the other. Timely vaccine updates in response to shifts in the viral fitness landscape may allow for more accurate targeting of emerging variants.

## Data Availability

ll data produced in the present study are available upon reasonable request to the authors

https://gisaid.org/

https://github.com/italia/covid19-opendata-vaccini

https://www.google.com/covid19/mobility/

https://covid19.apple.com/mobility

## Acknowledgments

We gratefully acknowledge all researchers for contributing to, generating, and openly sharing genome data via the Global Initiative on Sharing All Influenza Data (GISAID), as most of our analyses were based on this invaluable resource. We thank Prof. Haogao Gu and Dr. Jingbo Liang for helpful discussions, Ms. Fatema Khairunnasa for useful editing suggestions. We greatly appreciate Prof. Joshua S. Weitz at University of Maryland for his meaningful suggestions on model fitting. The authors also acknowledge support from grants funded by the [City University of Hong Kong] under Grant [#9610416]; [General Research Fund] under Grant [#9043514].

## Author Contributions

Zhaojun Ding and Hsiang-Yu Yuan designed the study. Zhaojun Ding collected, analyzed, and modelled the data. Zhaojun Ding and Hsiang-Yu Yuan wrote the paper. Hsiang-Yu Yuan gave final approval for publication. All authors read and approved the final manuscript.

## Data and code availability

Daily reported COVID-19 cases in Italy were collected from ‘Our World in Data’ (https://github.com/owid/covid-19-data/tree/master/public/data). Deep mutational scanning data were collected from “SARS-CoV-2 RBD DMS” (https://jbloomlab.github.io/SARS-CoV-2-RBD_DMS/). COVID-19 vaccination data in Italy were collected from the Italian Ministry of Health (https://github.com/italia/covid19-opendata-vaccini). Mobility data were collected from “Google COVID-19 Community Mobility Reports” (https://www.google.com/covid19/mobility/) and “Apple Map Mobility Trends Reports” (https://covid19.apple.com/mobility), respectively. The codes for this study are available at https://github.com/DING219/bind_escape_code.

## Ethics approval

This study is based on modeling using aggregate data, with no access to or analysis of individual-level information. Therefore, the study does not involve human participants and is not human subjects research. Ethical review and approval were not applicable.

