## Supplementary Material for "The role of receptor binding and immunity in SARS-CoV-2 fitness landscape: a modeling study"

**1. Supplementary data**

**1.1 VOCs samples collection**

We firstly downloaded the amino acid sequence data according to the classification options of each VOCs in the GISAID. Each sequence sample was including the information of collection date and amino acid sequence. Totally, after excluding the sequence with low quality according to the exclusion criteria mentioned in the main text, we made a summary statistic about them. The count of alpha, beta, delta, gamma and omicron sequences download from GISAID were 25155, 135, 41333, 2324 and 43064 through the whole study period, respectively. The proportion of beta and gamma are very low, only 2.07% and 0.12% (Fig. S1).

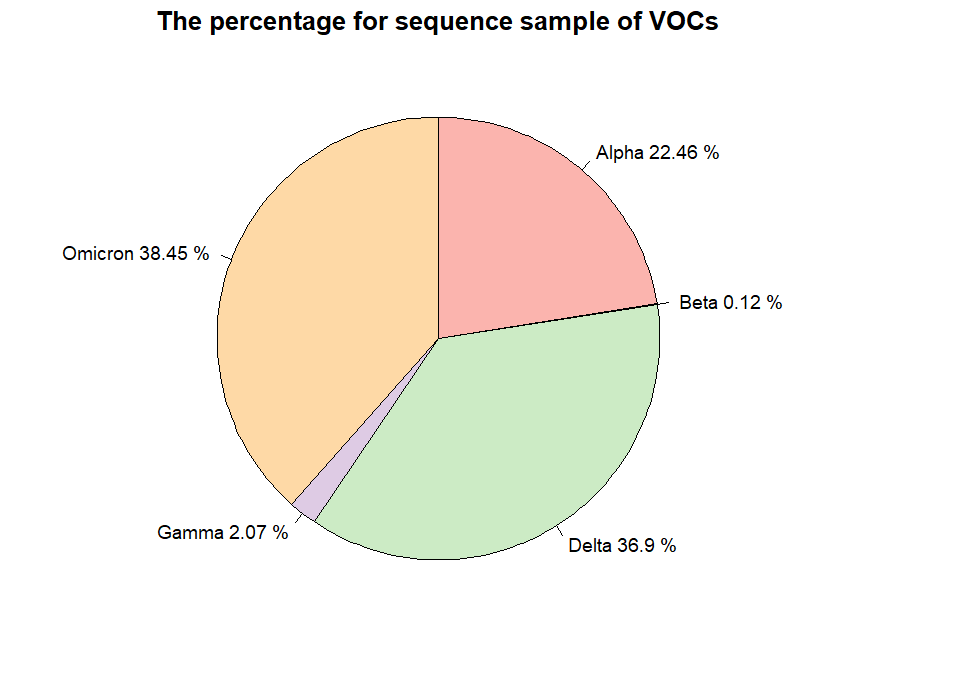

**Fig. S1. The percentage for samples of VOC sequence.** These data were selected by the variant option in the GISAID and downloaded in Oct-28, 2022. Each VOC sample was retrieved from GISAID website using the key words: “VOC Omicron (B.1.1.529+BA.*)”, “VOC Delta GK(B.1.617.2+AY.*)”, “VOC Alpha GRY(B.1.1.7+Q.*)”, “VOC Beta GH/501Y.V2(B.1.351+B.1.351.2+B.1.351.3)” and “VOC Gamma GR/501Y.V3(P.1+P.1.*)”.

**1.2 The relationship between daily incidence and mobility**

There is a significant negative correlation between mobility and daily incidence, indicating that as mobility increases, daily incidence decreases for Non-VOC, alpha and omicron strains. However, for the Delta variant, there is a positive correlation between mobility and daily incidence, implying that higher mobility is associated with an increase in daily cases. Even with significant correlations, the small absolute value of R coefficients suggests that while mobility may influence daily incidence, it does not fully explain the variation in daily incidence. Other factors, like social distancing and public awareness may play a role in determining virus transmission dynamics ^1^.

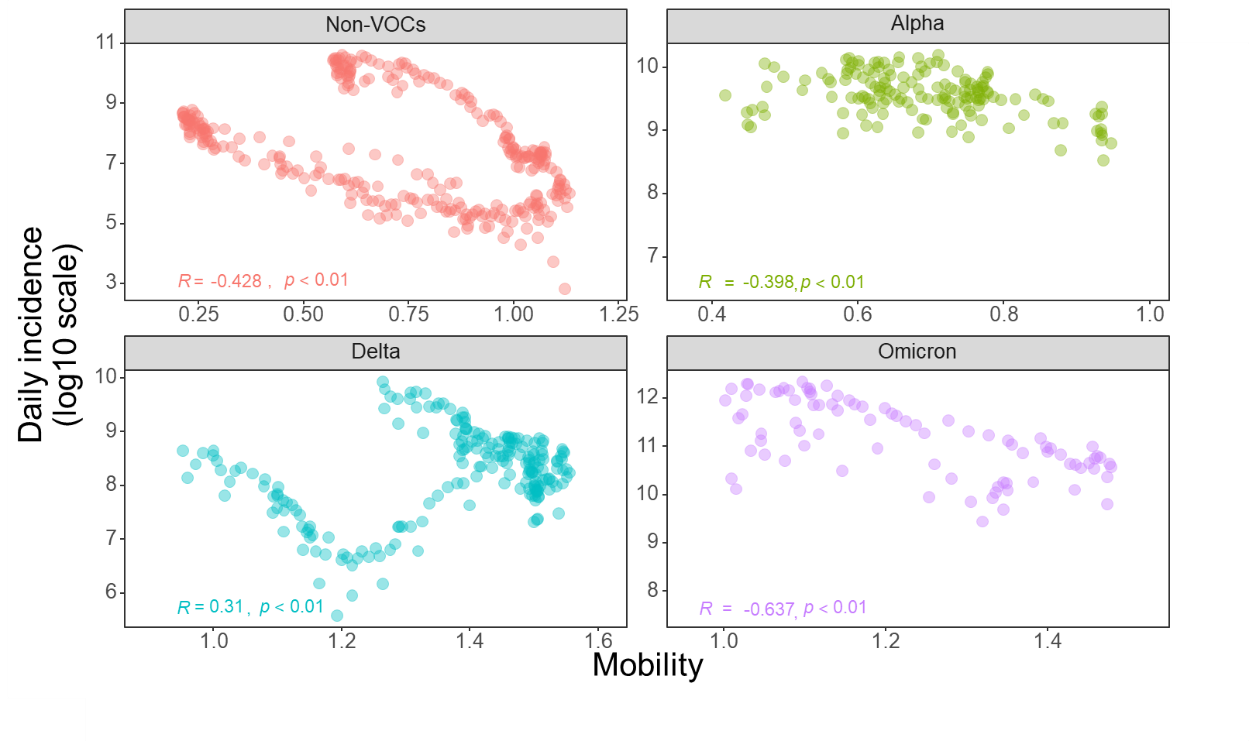

**Fig. S2. Correlation between mobility and daily COVID-19 incidence across SARS-CoV-2 variants (non-VOCs, Alpha, Delta, and Omicron).** The variants represented include non-VOCs, Alpha, Delta, and Omicron. Each plot displays the Pearson correlation coefficient (R) and the corresponding P-value.

**1.3 COVID-19 vaccination data in Italy**

The website (https://github.com/italia/covid19-opendata-vaccini) provided open data pertaining to the distribution and administration of COVID-19 vaccines across various regions in Italy. Daily population statistics for individuals who have received 1, 2, 3 or 4 doses were collected from Italy^2^. Vaccine manufacturers listed include Janssen, Moderna, Novavax, Pfizer Pediatrico, Pfizer/BioNTech, and Vaxzevria (AstraZeneca). For each manufacturer, the number of doses administered for each dose sequence is quantified (Table S1). Notably, Moderna and Pfizer/BioNTech show a significant number of doses administered across all categories, including booster doses, reflecting a continued vaccine uptake and possibly a booster campaign. The pie chart detailing the proportional distribution of different mRNA vaccines administered. Pfizer/BioNTech accounts for the majority at 63.3%, followed by Moderna at 24.7%, AstraZeneca at 9.04%, Pfizer Pediatric at 1.84%, and Novavax at 0.01%.

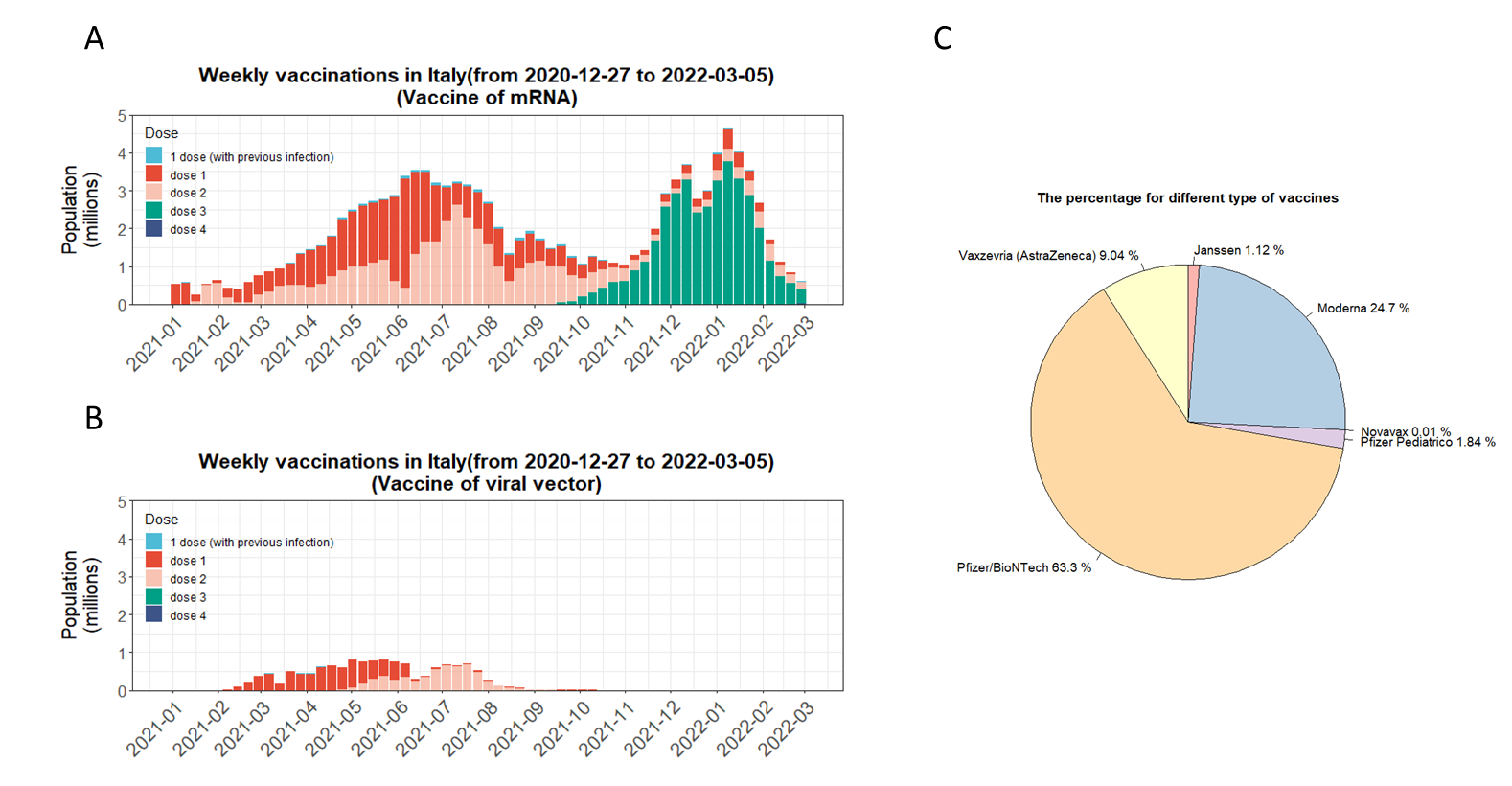

**Fig. S3. Vaccination trends and type distribution in Italy.** (A) mRNA vaccine administration timeline. (B) Viral vector vaccine administration timeline. (C) Proportional distribution for each type of vaccine.

**Table S1. The count of COVID-19 vaccination doses provided by each manufacturer.**

| **Vaccine manufacturers** | **d1** | **d2** | **dpi** | **db1** | **db2** | **total** |
| --- | --- | --- | --- | --- | --- | --- |
| Janssen | 1508016 | 0 | 0 | 0 | 0 | 1508016 |
| Moderna | 6661507 | 6641471 | 398692 | 19537150 | 4475 | 33243295 |
| Novavax | 9965 | 0 | 819 | 0 | 0 | 10784 |
| Pfizer Pediatrico | 1282120 | 1110032 | 77923 | 1891 | 0 | 2471966 |
| Pfizer/BioNTech | 32865745 | 32678265 | 1360533 | 18289241 | 10144 | 85203928 |
| Vaxzevria (AstraZeneca) | 6365179 | 5642377 | 165586 | 0 | 0 | 12173142 |

**1.4 Dynamics of virus binding and immune escape**

To illustrate the temporal dynamics of virus binding and immune escape more clearly, we employed a 1-week moving average method for the calculation of both binding and immune escape. The Fig. S4 provided a temporal analysis spanning from January, 2020, to May, 2022, charting the fluctuations in virus binding (depicted by the red line) and immune escape (illustrated by the blue line).

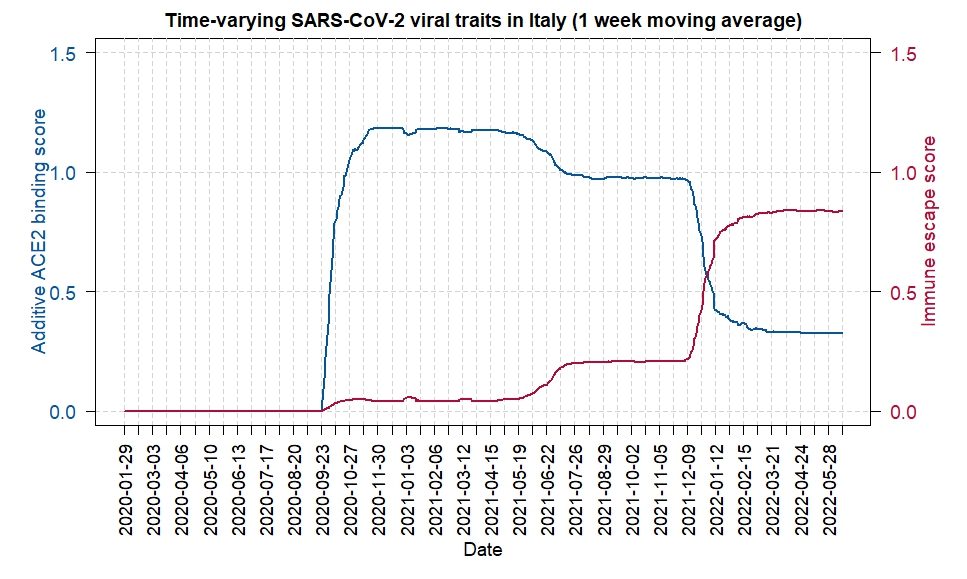

**Fig. S4. Temporal dynamics of virus binding and immune escape.** Both two virus traits (i.e. binding to ACE2 and immune escape) were calculated using a 1-week moving average to smooth short-term fluctuations and highlight longer-term trends.

**1.5** **Impact of mutations on ACE2 binding and immune escape across SARS-CoV-2 Variants**

Mutations (N501Y and E484K) generated notable increases in ACE2 additive binding, with N501Y showing a peak increase of approximately 0.25 in relative binding affinity, aligned with its significant prevalence across certain variants (Fig. S7A, B). This highlighted the substantial role that this mutation played in enhancing viral infectivity and transmissibility, consistent with previous studies. T478K and L452R prevalence was significantly higher in the delta variant (Fig. S7A), highlighting their contribution to heightened transmissibility and potential resistance to antibody neutralization when compared with earlier strains. Compared with previous VOCs, more mutations occurred in omicron, resulting in lower ACE2 binding (e.g. S371F, S375F, T376A, K417N, E484A, Q493R, and Y505H) and higher immune escape (e.g. K417N, E484A, and Q493R) (Fig. S7A).

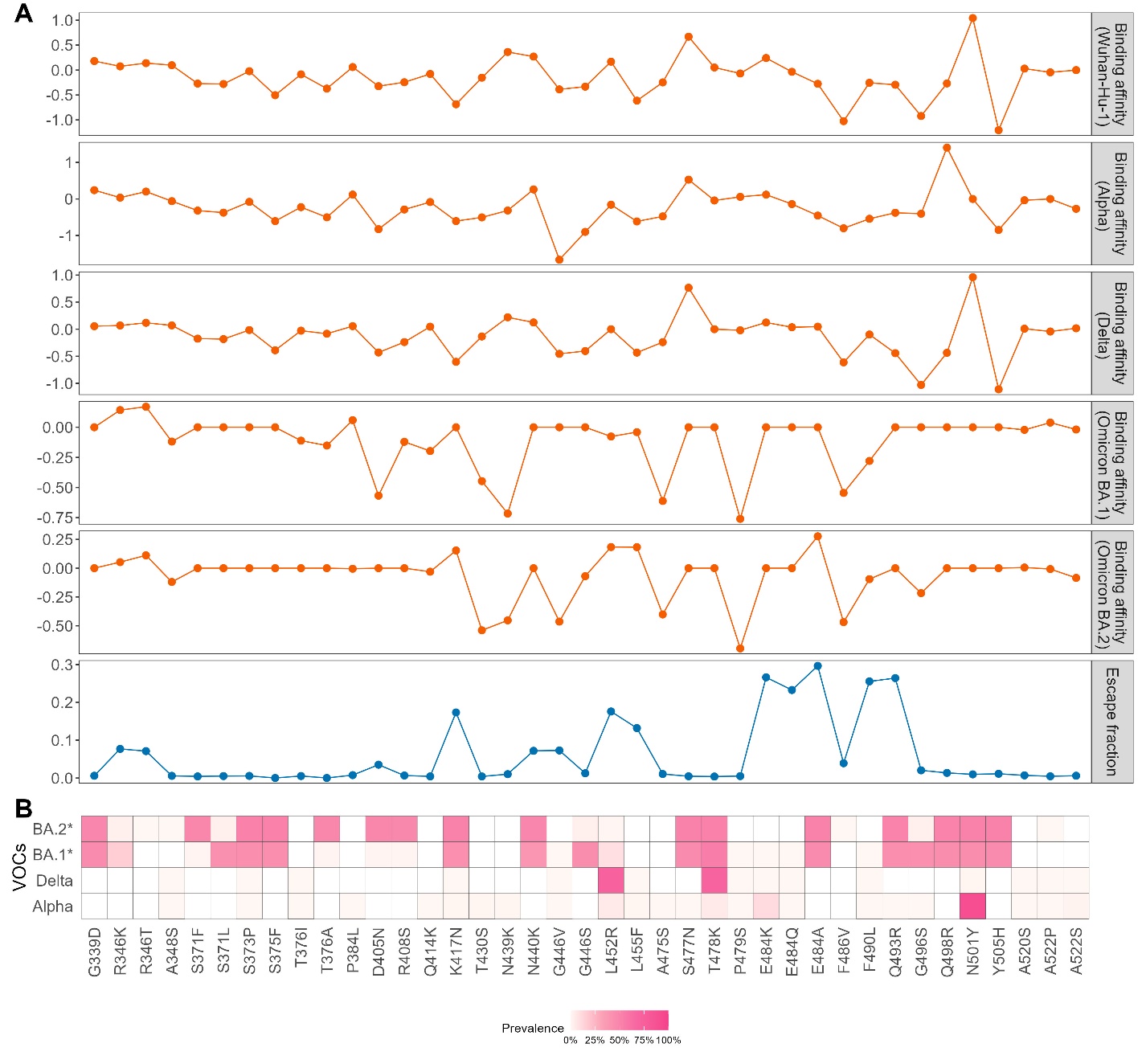

**Fig. S5. Mutations associated with different SARS-CoV-2 VOC and their effects on ACE2 binding affinity and immune escape.** (A) Mutation impact on ACE2 binding affinity and immune escape. The top graph shows the effects of specific mutations on spike protein binding affinity to the ACE2 receptor (referenced with multiple VOCs backgrounds), with values showing either increases or decreases in affinity. Note that binding scores of variant-defining mutations are denoted as 0, referring to the absence of those defining mutations related to the reference. The bottom graph depicts the same mutations’ impact on immune fraction, indicating the potential to escape antibody binding. (B) Mutation prevalence among each VOC. Heatmap showing specific mutation prevalence across alpha, delta, and omicron dominant periods.

**1.6 Time-varying prevalence of each mutations**

All mutations listed in the main text were showed here. It presents time series plots showing the prevalence of specific mutations in the RBD of SARS-CoV-2 spike protein over time, across different VOC. Some mutations are specific to certain variants (e.g., L452R and T478K in Delta), while others (e.g., N501Y) show up in multiple variants. The omicron variant introduces several new mutations (e.g., Q493R, N440K) that become prevalent during this period.

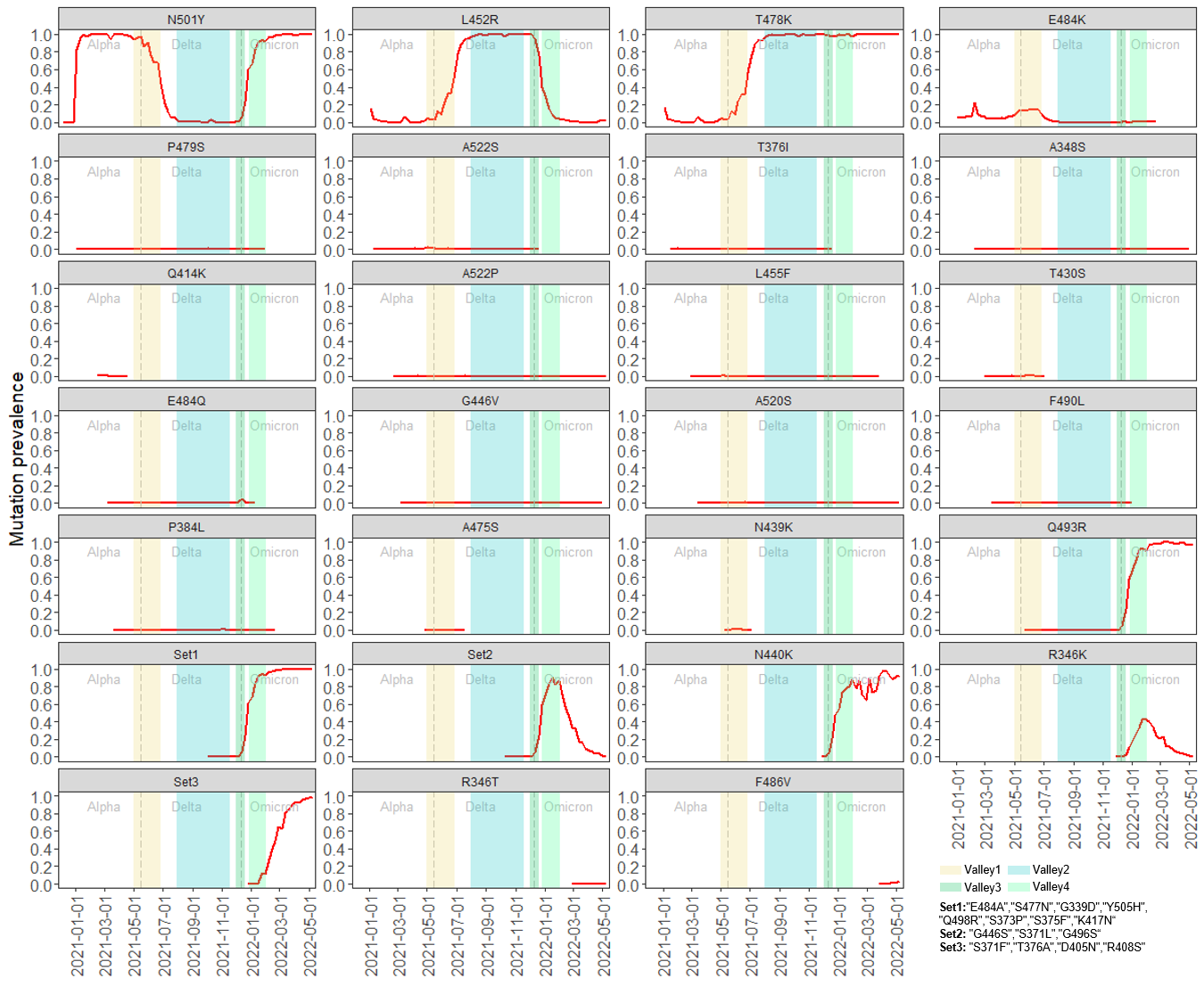

**Fig. S6. Mutation prevalence associated with VOC transitions.** Each plot tracks the prevalence of individual mutations (y-axis) over time (x-axis). Shadow regions corresponding to the valleys in main text Fig. 5D were marked by different colors. Some cluster mutations which have almost same prevalence were named as “Set 1”, “Set2” and “Set 3”. There are almost no mutations in RBD during the terminal of non-VOC period, therefore we don't consider the effect of mutations during non-VOC to Alpha.

**1.7 Sensitivity analysis about moderate vaccine waning scenario and fast waning scenario**

Several studies have examined the level of protection against reinfection with SARS-CoV-2, with an unweighted average of these studies indicating approximately 85.74% protection after 27.76 weeks ^3^. This finding aligns with the studies by Barnard et al., which also observed a gradual decline in immunity over time. In our study, this waning rate, represented by log(0.85/182.5), was applied to both vaccine-induced protection and immunity resulting from natural infection ^4^.

To account for potential variability in the rate of immunity loss, alternative waning scenarios were considered. One scenario assumes a moderate decline in vaccine protection, with a 50% loss of protection after 6 months (log(0.5/182.5)) (Fig. S8A), while a faster waning scenario assumes a 50% loss of protection after 3 months (log(0.5/90)) (Fig. S8B). When comparing the results from these moderate and faster waning scenarios with the optimistic scenario (log(0.85/182.5)), the overall viral evolution pattern keep similarly. Despite the different assumptions about vaccine waning rates, the relationship between ACE2 binding score, effective immunity, and fitness follows similar trends, indicating that the rate of vaccine protection waning does not significantly affect the conclusions regarding viral evolution.

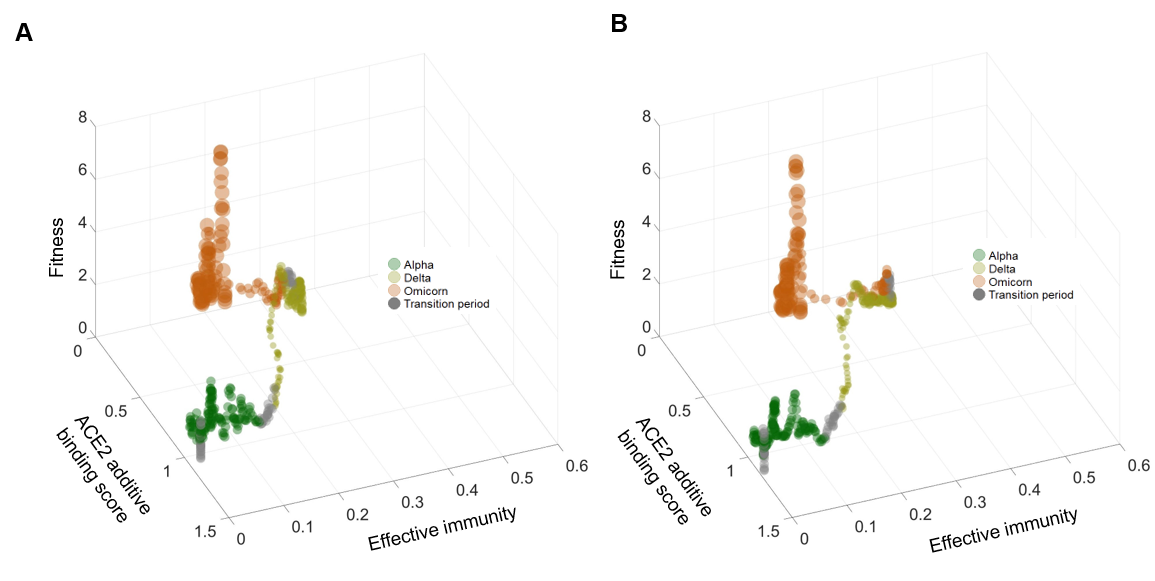

**Fig. S7. Interplay between ACE2 additive binding score, effective immunity, and virus’s fitness in SARS-CoV-2 evolution.** (A) The moderate vaccine waning scenario, where protection decreases by 50% after 6 months (log(0.5/182.5)). (B) The faster waning scenario, with 50% protection loss after 3 months (log(0.5/90)).

**2. Supplementary methods**

**2.1 Binding and immune escape measurement**

The deep mutational scanning data ^5^ and escape fraction data ^6^for SARS-CoV-2 RBD mutations were utilized to calculate the virus binding and immune escape, respectively. The calculation process was shown in Fig. S10. For each amino acid sequence, all positions will be allocated the score that measures the binding affinity to ACE2 and escape fraction of antibody binding. Virus total binding and immune escape were calculated by adding the value together for each sequence, which contribute to quickly calculating^7^.

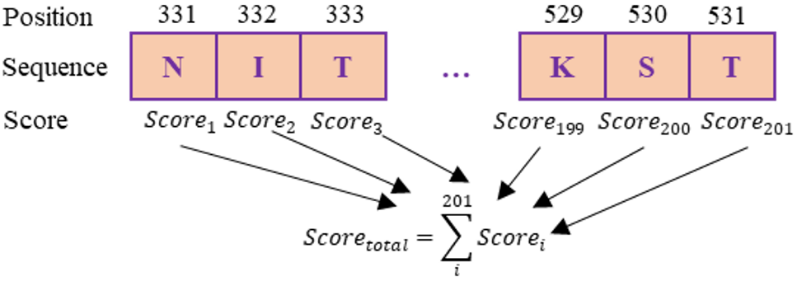

**Fig. S8. ACE2 binding and immune escape calculation process.** The number above represents the site of the amino acid sequence of the spike protein, RBD region (residues from N331 to T531).

**2.2 Model details**

**2.2.1 Modelling transmission dynamics**

Individuals were classified into the following infection classes (main text Fig. 1): susceptible ( $S_{i}$), exposed (but not yet infectious, $E_{i}$), infectious ($I_{i}$), recovered ($R_{i}$) and the confirmed cases by reporting ($\mathrm{IR}$). In addition, we also introduced the daily number of vaccinated people with 1,2 and 3 doses into the model to calibrate the changes of susceptible people ($V_{i}$). The key epidemiological and model-specific parameters, including fixed values, estimated quantities, and assumed constants, that were used in the transmission dynamics framework were summarized in Table S2. $\beta_{imm\_i}(t)$ is the transmission rate between infectious and people in each susceptible level; $\sigma$ is the reverse of incubation period, which was set as 1/5.5 ^8,9^; $\rho_{t}$ is the confirmation delay, which was set as 1/3 ^10^; $d$ is the detection rate, which was estimated as 0.82 (95%CI: 0.71-0.94) from model fitting; $\omega$ is the rate of waning immunity for the recovered people, which was set as $log(0.85)/182.5$. The recovery rate, denoted by $\gamma_{t}$, exhibits a piecewise linear form with changepoints determined based on the distinct VOC periods:

$$\gamma_{t}=\left\{ \begin{matrix} \begin{matrix} \gamma_{Non-VOC}, during Non-VOC period \\ \gamma_{Alpha}, during Alpha period \end{matrix} \\ \begin{matrix} \gamma_{Delta}, during Delta period \\ \gamma_{Omicron}, during Omicorn period \end{matrix} \end{matrix} \right.$$

Where$\gamma_{Non-VOC}=1/8.9$, $\gamma_{Alpha}=1/4.4$, $\gamma_{Delta}=1/5.2$ and $\gamma_{Omicron}=1/3$ according to the previous studies ^11,12^. The Non-VOC period, alpha period, delta period and omicorn period were defined from February-21, 2020 to February-4, 2021, February-5,2021 to June-23, 2021, June-24, 2021 to October-1, 2021 and October-2, 2021 to March-5, 2022, respectively ^8^. The model equations for the transmission process showed in main text Fig. 1 were write as follows:

$$\frac{dS_{0}}{dt}=\omega(R_{0}+R_{1}+R_{2}+R_{3}+S_{1}+S_{2}+S_{3})-\frac{\beta_{imm\_0}\left( t \right)\cdot\sum_{i=0}^{4} I_{i}\cdot S_{0}}{N}-V_{1}$$

$$\frac{dE_{0}}{dt}=\frac{\beta_{imm\_0}\left( t \right)\cdot\sum_{i=0}^{4} I_{i}\cdot S_{0}}{N}-\sigma\cdot E_{0}$$

$$\frac{dI_{0}}{dt}=\sigma\cdot E_{0}-\gamma_{t}\cdot I_{0}-\rho_{t}\cdot I_{0}$$

$$\frac{dR_{0}}{dt}=\gamma_{t}\cdot{IR}_{0}+\gamma_{t}\cdot I_{0}-\omega\cdot R_{0}$$

$$\frac{d{IR}_{0}}{dt}={d\cdot\rho}_{t}\cdot\sigma\cdot E_{0}$$

$$\frac{dS_{1}}{dt}=-\frac{\beta_{imm\_1}\left( t \right)\cdot\sum_{i=0}^{4} I_{i}\cdot S_{1}}{N}-\omega\cdot S_{1}+V_{1}-V_{2}$$

$$\frac{dE_{1}}{dt}=\frac{\beta_{imm\_1}\left( t \right)\cdot\sum_{i=0}^{4} I_{i}\cdot S_{1}}{N}-\sigma\cdot E_{1}$$

$$\frac{dI_{1}}{dt}=\sigma\cdot E_{1}-\gamma_{t}\cdot I_{1}-\rho_{t}\cdot I_{1}$$

$$\frac{dR_{1}}{dt}=\gamma_{t}\cdot{IR}_{1}+\gamma_{t}\cdot I_{1}-\omega\cdot R_{1}$$

$$\frac{d{IR}_{1}}{dt}={d\cdot\rho}_{t}\cdot\sigma\cdot E_{1}$$

$$\frac{dS_{2}}{dt}=-\frac{\beta_{imm\_2}\left( t \right)\cdot\sum_{i=0}^{4} I_{i}\cdot S_{2}}{N}-\omega\cdot S_{2}+V_{2}-V_{3}$$

$$\frac{dE_{2}}{dt}=\frac{\beta_{imm\_2}\left( t \right)\cdot\sum_{i=0}^{4} I_{i}\cdot S_{2}}{N}-\sigma\cdot E_{2}$$

$$\frac{dI_{2}}{dt}=\sigma\cdot E_{2}-\gamma_{t}\cdot I_{2}-r\cdot\rho_{t}\cdot I_{2}$$

$$\frac{dR_{2}}{dt}=\gamma_{t}\cdot{IR}_{2}+\gamma_{t}\cdot I_{2}-\omega\cdot R_{2}$$

$$\frac{d{IR}_{2}}{dt}={d\cdot\rho}_{t}\cdot\sigma\cdot E_{2}$$

$$\frac{dS_{3}}{dt}=-\frac{\beta_{imm\_3}\left( t \right)\cdot\sum_{i=0}^{4} I_{i}\cdot S_{3}}{N}-\omega\cdot S_{3}+V_{3}$$

$$\frac{dE_{3}}{dt}=\frac{\beta_{imm\_3}\left( t \right)\cdot\sum_{i=0}^{4} I_{i}\cdot S_{3}}{N}-\sigma\cdot E_{3}$$

$$\frac{dI_{3}}{dt}=\sigma\cdot E_{3}-\gamma_{t}\cdot I_{3}-r\cdot\rho_{t}\cdot I_{3}$$

$$\frac{dR_{3}}{dt}=\gamma_{t}\cdot{IR}_{3}+\gamma_{t}\cdot I_{3}-\omega\cdot R_{3}$$

$$\frac{d{IR}_{3}}{dt}={d\cdot\rho}_{t}\cdot\sigma\cdot E_{3}$$

**2.2.2 Virus transmission rate and fitness**

To explore the effect of virus intrinsic factors (i.e. binding and immune escape) and extrinsic factors (i.e. population immunity) on the virus transmissibility, the virus transmission rate can be calculated like follows:

$$\beta_{imm\_i}\left( t \right)=\beta_{v}\left( t \right)\cdot(1-V_{e_{i}}\cdot e^{-c\cdot immune\_escape\_t})\cdot M(t)$$

The transmission rate, $\beta_{v}\left( t \right)$, which was influenced by the intrinsic factors of virus, was defined as the virus fitness. $c$ is the scaling factor, which was estimated as 0.67 (95%CI: -1.24-2.92) from model fitting. And it would be evaluated followed Brownian motion^13^. The Brownian motion factor was assumed to follow the normal distribution with the standard deviation (SD) parameter as 0.5:

$$\tau\sim\mathrm{Normal}\left( 0, 0.5 \right)$$

$$log(\beta_{v}\left( t \right))={log(\beta}_{v}\left( t-1 \right))+\tau$$

All parameter specifications and estimation procedures for the infectious-disease transmission model are provided in Supplementary Table S2.

**Table S2. Summary of parameters used in the transmission dynamics model**

| **Parameter** | **Notation** | **(Value)/Range** | **Status** | **Source(s)** |
| --- | --- | --- | --- | --- |
| Transmission rate | β_v_(t) | Time-varying | to be estimated | – |
| Incubation period | σ⁻¹ | 5.5 | fixed | 8,9 |
| Confirmation delay | ρ_t_⁻¹ | 3 | fixed | 10 |
| Detection rate | d | 0.77 (95% CI: 0.55–0.94) | to be estimated | – |
| Waning immunity rate | ω | log(0.85)/182.5 | fixed | 4 |
| Recovery rate | γ_t_ | Piecewise linear | fixed | 11,12 |
| Scaling factor for viral imnue escape | c | 0.67 (95% CI: -1.24–2.92) | to be estimated | – |
| SD of brownian motion factor | σ | 0.5 | fixed | 10,13 |

**2.2.3** **Population immunity**

Herd immunity or population immunity is essentially a simple concept describing the totality of naturally acquired and vaccine-based immunity to a given infectious agent as a proportion of the whole population ^14,15^. It can be formulated with full protection as:

$$P_{imm\_total}\boldsymbol{=}\frac{Rco+Vac}{N}$$

Where $Rco$ is the count of recovered people from natural infection; $Vac$ is the count of vaccinated people (i.e. vaccine coverage of primary dose);$N$ is the whole population; Term $\frac{Vac}{N}$ was indicated as the vaccine-induced immunity. The population immunity calculation above assumed that the vaccine or recovering from infection is 100% protective. Here, we supposed that the protection of the people who recovered from previous infection, its protection can be denoted as $V_{r}$. The people who got vaccinated, its protection can be denoted as $V_{e}$. Considering the immunized individuals may experience waning immunity over time, resulting in a decrease in their level of protection, the protection can be rewrite as $V_{e_{t}}=V_{e_{0}}\cdot e^{-\lambda t}$ , $V_{r_{t}}=V_{r_{0}}\cdot e^{-\lambda t}$ at day $t$, where$V_{e_{0}}$and $V_{r_{0}}$ are the initial VE for the protection of people who got vaccine or previous infection, the values of $V_{e_{0}}$ for 1, 2 and 3 doses of mRNA vaccine are 0.79, 0.89 ^16^ and 0.95 ^17^, respectively and for viral vector vaccine (only up to 2 doses of viral vector vaccine administered for Italy population during our study period) are 0.55 and 0.65, respectively ^16^; $V_{r_{0}}$ was assumed to equal to initial VE of viral vector vaccine; the $\lambda=log(0.85)/182.5$, corresponding to exponential waning with a 15% loss of protection after 6 months ^4^. Next, with the definition of daily vaccination number or recovered number, the protection can be calculated as:

Day 1: $P_{imm\_total}\left( 1 \right)=(\sum_{k=0}^{N_{dose}} {Vac}_{1}^{k}V_{e_{1}}^{k}+{Rco}_{1}\cdot V_{r1})/N$

Day 2: $P_{imm\_total}\left( 2 \right)=(\sum_{k=0}^{N_{dose}} {Vac}_{2}^{k}V_{e_{1}}^{k}+{Rco}_{2}\cdot V_{r1}+\sum_{k=0}^{N_{dose}} {Vac}_{1}^{k}V_{e_{2}}^{k}+{Rco}_{1}\cdot V_{r2})/N$

Day3:

$$P_{imm\_total}\left( 3 \right)=(\sum_{k=0}^{N_{dose}} {Vac}_{3}^{k}V_{e_{1}}^{k}+{Rco}_{3}\cdot V_{r1}+\sum_{k=0}^{N_{dose}} {Vac}_{2}^{k}V_{e_{2}}^{k}+{Rco}_{2}\cdot V_{r2}+\sum_{k=0}^{N_{dose}} {Vac}_{1}^{k}V_{e_{3}}^{k}+{Rco}_{1}\cdot V_{r_{3}})/N$$

……..

Day t: $P_{imm\_total}\left( t \right)=(\sum_{t=1}^{N_{day}} (\sum_{k=0}^{N_{dose}} {Vac}_{t}^{k}V_{e_{N-\left( t-1 \right)}}^{k}+{Rco}_{t}*V_{r_{N-\left( t-1 \right)}}))/N$

Where $t$ stands for a single day during study period; $k$ means the 1, 2 or 3 doses vaccination people got. To adjust the overestimation of protection for the immunized people, we assumed that 2 or 3 doses are assumed to be administered to all individuals who have been vaccinated with 1 dose after an average of 42 days since the previous dose. So, the equation above can be adjusted as follows:

$$P_{imm\_total}\left( t \right)=\sum_{t=1}^{N_{day}} (\sum_{k=0}^{N_{dose}} m_{t}^{k}V_{e_{N-\left( t-1 \right)}}^{k}+{Rco}_{t}*V_{r_{N-\left( t-1 \right)}}-\sum_{k=0}^{N_{dose}} m_{t}^{k}V_{e_{N-\left( t-1 \right)-41}}^{k})/N$$

Similarly, if we only consider the vaccine induced population immunity (which means only the count of vaccinated people are used to calculate the $P_{imm}$), the protection (only induced by vaccine, which denoted as $P_{imm\_vac}$) can be calculated as:

$$P_{imm\_vac}\left( t \right)=\sum_{t=1}^{N_{day}} (\sum_{k=0}^{N_{dose}} m_{t}^{k}V_{e_{N-\left( t-1 \right)}}^{k}-\sum_{k=0}^{N_{dose}} m_{t}^{k}V_{e_{N-\left( t-1 \right)-41}}^{k})/N$$

**2.2.4 Reconstruction of the** **fitness landscape**

To reconstruct the rugged fitness landscape, we adopted a Gaussian Mixture Model (GMM). Five peaks were assumed to represent the major variant clusters, with the mean values of the multivariate Gaussian components chosen according to the observed daily average ACE2 binding and effective immunity coordinates of the corresponding variants of concern (VOCs). Covariance matrices for each Gaussian were specified to reflect the empirical variability in binding–immunity space, with some peaks modeled to have elongated shapes along the direction of the observed evolutionary trajectories (i.e., anisotropic covariance aligned with the line connecting adjacent peaks).

Synthetic samples were generated from these five Gaussian distributions with weights proportional to the relative prominence of each peak. A GMM with five full covariance components was then fitted to the combined sample using the Expectation–Maximization algorithm (fitgmdist function in MATLAB (R2023b)), with 10 replicates, a regularization term to ensure numerical stability, and a maximum of 2000 iterations to ensure convergence.

For visualization, the fitted probability density function (PDF) was evaluated on a two-dimensional grid covering binding (0–1.5) and effective immunity (0–0.6). The resulting PDF values were normalized and rescaled so that the maximum height corresponded to 8 units of fitness, consistent with the scale used in the main text. To further smooth the surface and enhance continuity between peaks, the grid was convolved with a Gaussian kernel (window size 11, standard deviation 1.4). This smoothing procedure is equivalent to kernel density smoothing, where the kernel bandwidth is explicitly defined by the kernel size and variance parameters. Finally, the landscape was rendered as a three-dimensional surface (surfl function with interpolated shading). Results were visually and numerically robust to (1) moderate changes in kernel width (range from 1.0 to 1.8]), (2) halving/doubling grid resolution, and (3) alternative EM initializations (top-2 likelihood solutions among 10 replicates were nearly indistinguishable).
